# Utilizing Deep Learning to Improve Diagnostic Accuracy in Glioma, Pituitary Tumors, and Meningiomas

**DOI:** 10.1101/2024.12.10.24318709

**Authors:** Alon Gorenshtein, Tom Liba, Avner Goren

## Abstract

**Background:** Glioma, pituitary tumors, and meningiomas constitute the major types of primary brain tumors. The challenge in achieving a definitive diagnosis stems from the brain’s complex structure, limited accessibility for precise imaging, and the resemblance between different types of tumors. An alternative and promising solution is the application of artificial intelligence (AI), specifically through deep learning models.

**Methods:** This study developed a brain tumor detection model using a comprehensive dataset of 7,023 MRI images. Included 1,621 images for glioma, 1,645 for meningioma, 1,757 for pituitary tumors, and 2,000 for non-tumor magnetic resonance imaging (MRI) images. We employed deep learning, specifically convolutional neural network (CNN). We utilized the ResNet-18 architecture, a widely used pre-trained model. We fine-tuned the pre-trained ResNet-18 on our dataset, which included MRI images classified into glioma, pituitary, and meningioma tumors. The fine-tuning was conducted over 10 epochs. After fine-tuning, the model that demonstrated the highest performance on validation data was selected as the best model

**Results:** The deep learning model demonstrated exceptional performance across all diagnostic categories, with the training accuracy stabilizing at nearly 100% and validation accuracy closely mirroring this trend. The model achieved an AUC of 1.00 across all categories, with sensitivities of 95% for glioma, 98% for meningoma, 99% for pituitary tumors, and 100% for non-tumor MRI images.

**Conclusion:** Our study utilized a fine-tuned Residual Network 18 layers (ResNet-18) model in PyTorch to significantly enhance the diagnostic accuracy of primary brain tumors, specifically gliomas, meningiomas, and pituitary tumors. The model demonstrated high accuracy, indicating its potential to improve early diagnosis and patient outcomes. These findings suggest that integrating AI models like ResNet-18 into clinical workflows could lead to earlier detection and more precise prognostic evaluations, ultimately enhancing patient care and survival rates.

## Introduction

Glioma, meningiomas, and pituitary tumors constitute the major types of primary brain tumors, accounting for 24.5%, 39%, and 17.1% respectively [1]. Magnetic resonance imaging (MRI) with gadolinium enhancement is the preferred diagnostic tool for these conditions [2]. Standard imaging protocols in neuro-oncology, including T1-weighted, T2-weighted, T2-weighted fluid-attenuated inversion recovery (FLAIR), and post-contrast T1-weighted sequences, have demonstrated good accuracy in the diagnosis of primary brain tumors [3].

While pituitary tumors and meningiomas typically present benign characteristics, gliomas often manifest more aggressively and account for 75% of malignant primary brain tumors in adults. Only 1% of gliomas are non-malignant, underscoring the critical need to differentiate between gliomas and benign tumor types [1].

Despite the usage of the magnetic resonance imaging (MRI) technology, definitive diagnosis remains challenging due to the brain’s complex anatomy, restricted access for precise imaging [4], and similarities among different tumor types. To address the diagnostic challenges posed by brain tumors, several approaches have been considered. Advancements in standard MRI techniques such as gadolinium enhancement have been supplemented by more sophisticated imaging methods including diffusion-weighted MRI, perfusion-weighted MRI, MR spectroscopy, and positron emission tomography [5]. However, these advanced techniques are not commonly utilized in routine practice and often require specialized interpretation by professionals.

An alternative and promising solution is the application of artificial intelligence (AI), specifically through deep learning models. These models are extensively trained to enhance their accuracy and are fine-tuned to generalize learned patterns, enabling them to effectively differentiate between tumorous, and non-tumorous MRI images [6]. The overarching goal of this study is to develop a deep learning model capable of diagnosing MRI images to determine whether they depict one of the three tumor types or are non-tumorous, potentially transforming neuro-oncology by providing precise and automated diagnoses.

## Materials and Methods

### Ethical Compliance

The present study was deemed exempt from Institutional Review Board (IRB) approval as it only utilized a publicly available dataset and did not involve human subjects. The dataset employed for this research was gathered from openly accessible sources, ensuring complete anonymity and confidentiality of any personal information. Therefore, no ethical concerns regarding human subject involvement were applicable to this investigation, allowing us to proceed without the need for IRB approval.

### Dataset

This study developed a brain tumor detection model using a comprehensive dataset of 7,023 MRI images sourced from Kaggle.com [7]. The dataset, compiled by Nickparvar by merging three smaller datasets, included 1,621 images for glioma, 1,645 for meningioma, 1,757 for pituitary tumors, and 2,000 for non-tumor MRI images. The goal was to train a model to recognize specific patterns indicative of tumors, thus enabling automated and accurate detection. The dataset was curated to reflect a diverse range of tumors in terms of grade, type, and presentation, enhancing the model’s applicability across different clinical settings. The images, obtained from various imaging devices via Kaggle.com, contribute to the model’s generalizability.

### Data Characteristics

The MRI images used in this study varied in both orientation (horizontal, sagittal) and type (T1, T2), encompassing a wide range of anatomical perspectives and magnetic properties. This variability ensures the model is not biased towards any specific orientation or imaging parameter, enhancing its generalizability across different clinical settings and machine configurations.

### Deep Learning Approach

In this study, we employed deep learning due to its demonstrated efficacy in medical image analysis, particularly for recognizing complex patterns in imaging data. We selected a convolutional neural network (CNN), a type of deep learning model particularly adept at processing images. CNNs automate the feature extraction process, reducing the need for manual intervention and allowing for more robust data analysis.

### Model Selection and Implementation

We utilized the ResNet-18 architecture, a widely used pre-trained model known for its ability to train deeper networks by using residual connections to prevent the vanishing gradient problem. This choice was influenced by ResNet-18’s success in various image classification tasks and its suitability for our dataset size and computational limits. We fine-tuned the pre-trained Residual Network 18 layers (ResNet-18) on our dataset, which included MRI images classified into glioma, pituitary, and meningioma tumors. The fine-tuning was conducted over 10 epochs to adapt the model to the specific features of brain tumors while leveraging the general image recognition capabilities learned from the larger ImageNet dataset. After fine-tuning, the model that demonstrated the highest performance on validation data was selected as the best model for further evaluation. This structured approach allowed the model to learn from a substantial training set of 5,712 images, tested against 1,311 images to ensure robustness and accuracy. Figure 1. Depict the labeling of each tumor and non-tumor MRI.

**Figure 1.**
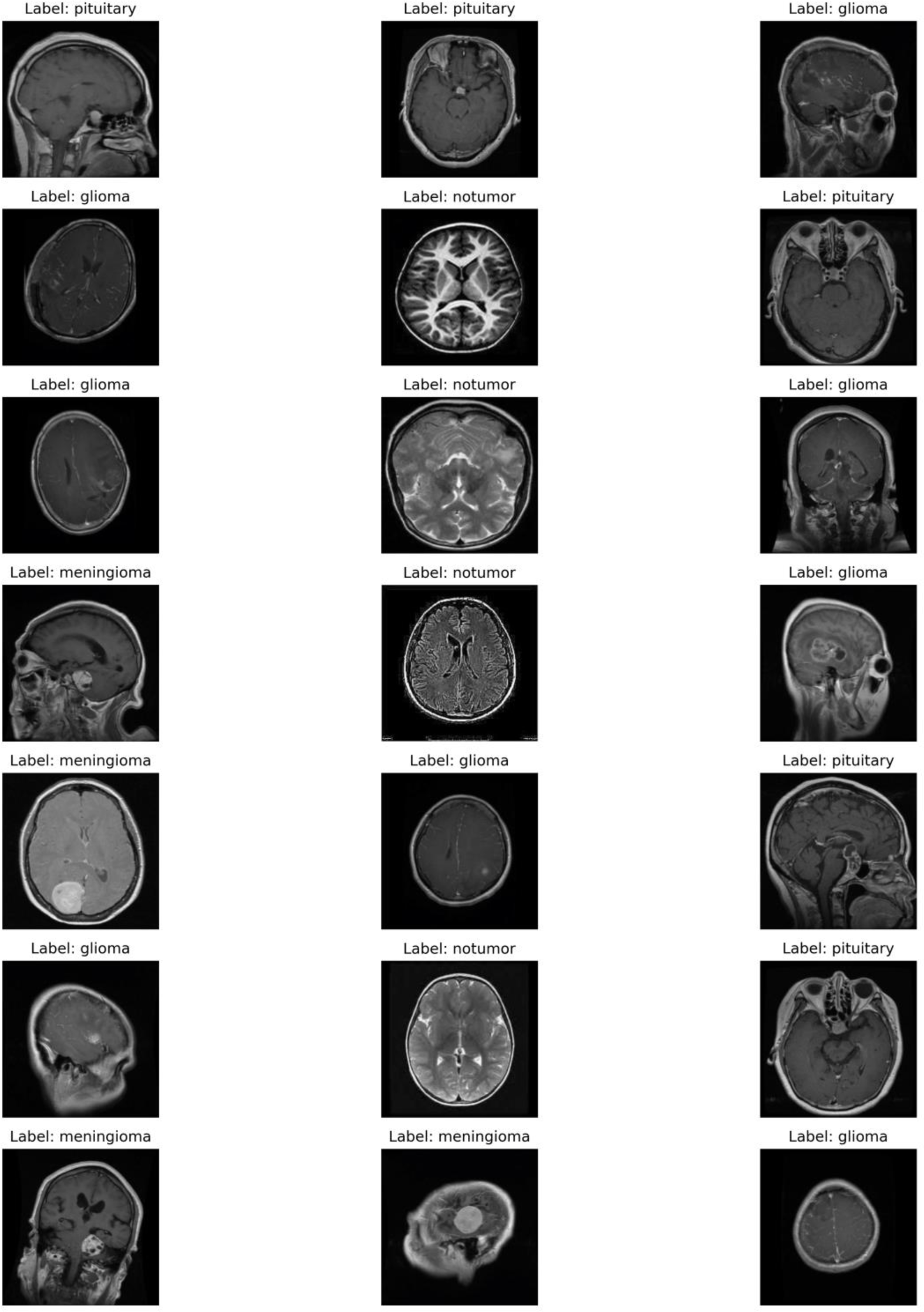
the labeling of the MRI images used in this study varied in both orientation (horizontal, sagittal) and type (T1, T2).

### Model development

The model development was facilitated by Google’s collaboration platform, which supported efficient data preprocessing, feature extraction, and model training phases.

### Model performance assessments

The model’s performance was rigorously assessed through several statistical metrics: accuracy, precision, recall, sensitivity, specificity, and the F1 score. These metrics were crucial for gauging the model’s proficiency in correctly identifying tumor-positive cases, as well as its effectiveness in minimizing both false positives and false negatives. This comprehensive analysis provided critical insights into the model’s diagnostic accuracy and potential clinical applicability.

## Results

### Model performance

The deep learning model demonstrated exceptional performance across all diagnostic categories. The training and validation accuracy remained consistently high throughout the training process, with the training accuracy stabilizing at nearly 100% and validation accuracy closely mirroring this trend (Figure 2). This high level of accuracy in both the training and validation phases indicates a robust model with excellent generalization capabilities.

**Figure 2.**
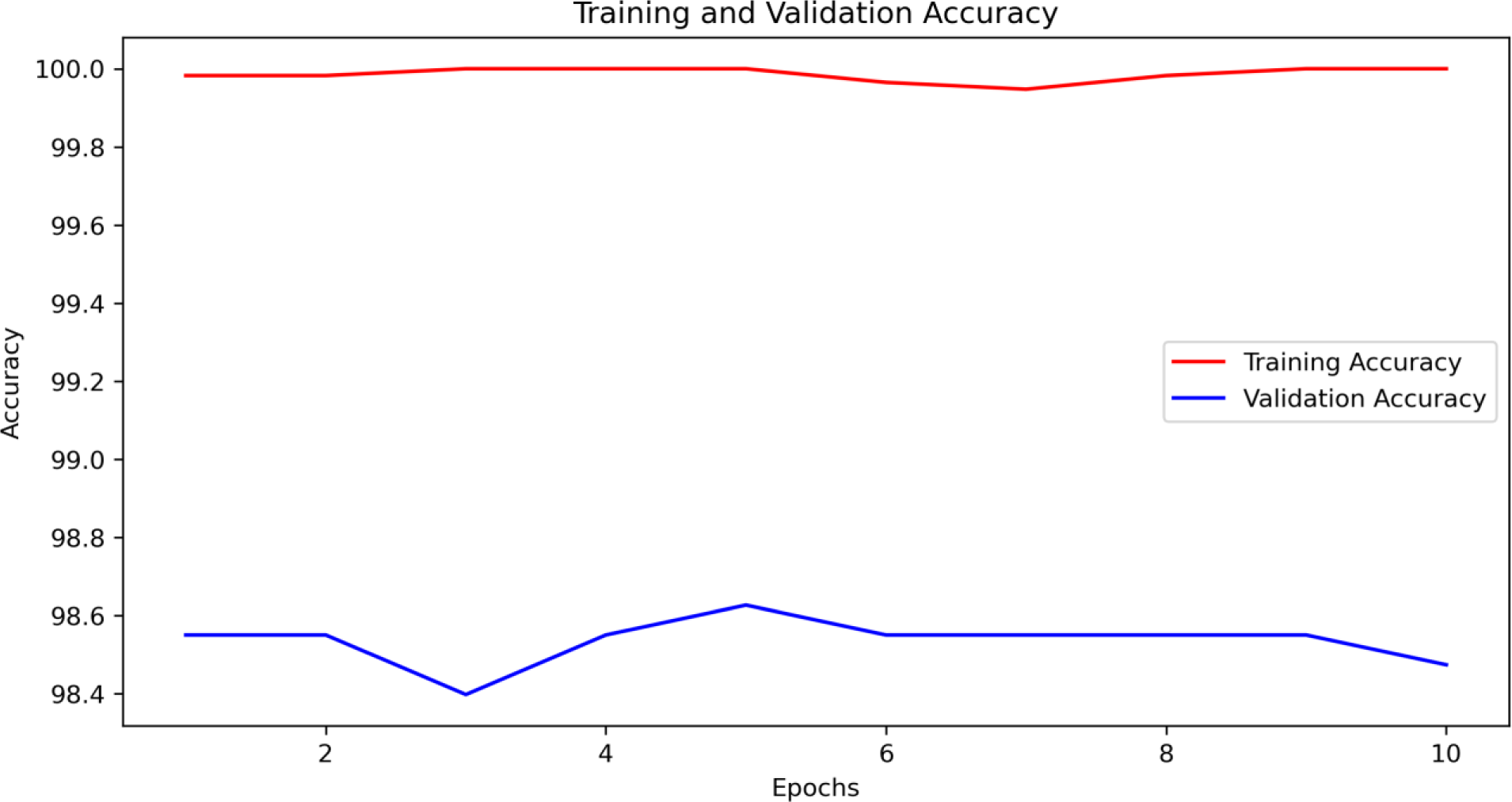
The training and validation accuracy curves of the ResNet-18 model fine-tuned for diagnosing gliomas, meningiomas, and pituitary tumors.

### Diagnostic Ability

The Receiver Operating Characteristic (ROC) curves for each diagnostic category (glioma, meningioma, no tumor, and pituitary tumor) further illustrate the model’s diagnostic prowess (Figure 3). All categories achieved an area under the curve (AUC) of 1.00, indicating perfect discrimination between positive and negative cases for each category. Table 1 showcases the diagnostic performance metrics.

**Figure 3.**
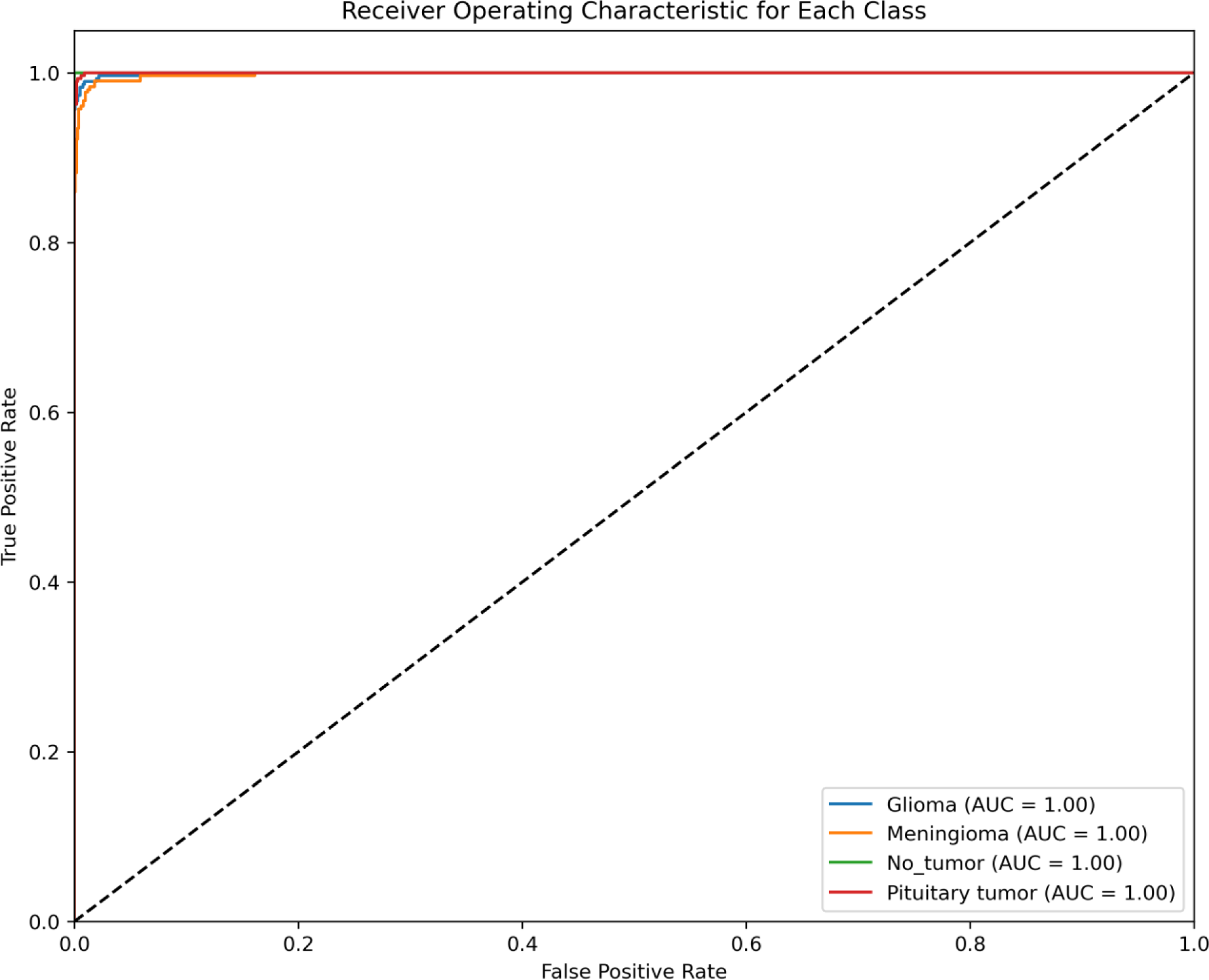
Area Under the Curve (AUC) for each type of brain tumor diagnosis (gliomas, meningiomas, and pituitary tumors).

**Table 1.** Performance metrics for each diagnosis.

| Diagnosis | Sensitivity | Specificity | PPV | NPV | AUC |
| --- | --- | --- | --- | --- | --- |
| Glioma | 0.95 | 0.99 | 0.99 | 0.98 | 1 |
| Meningioma | 0.98 | 0.98 | 0.96 | 0.99 | 1 |
| Pituitary tumor | 0.99 | 0.99 | 0.98 | 0.99 | 1 |
| No tumor | 1 | 0.99 | 0.99 | 1 | 1 |

### Error Analysis

The confusion matrix provided a detailed view of the model’s predictive accuracy across different classes (Figure 4). The matrix showed predominant correct classifications with minimal confusion between classes, demonstrating the model’s precise discriminatory capability between different tumor types and non-tumor types of MRI images.

**Figure 4.**
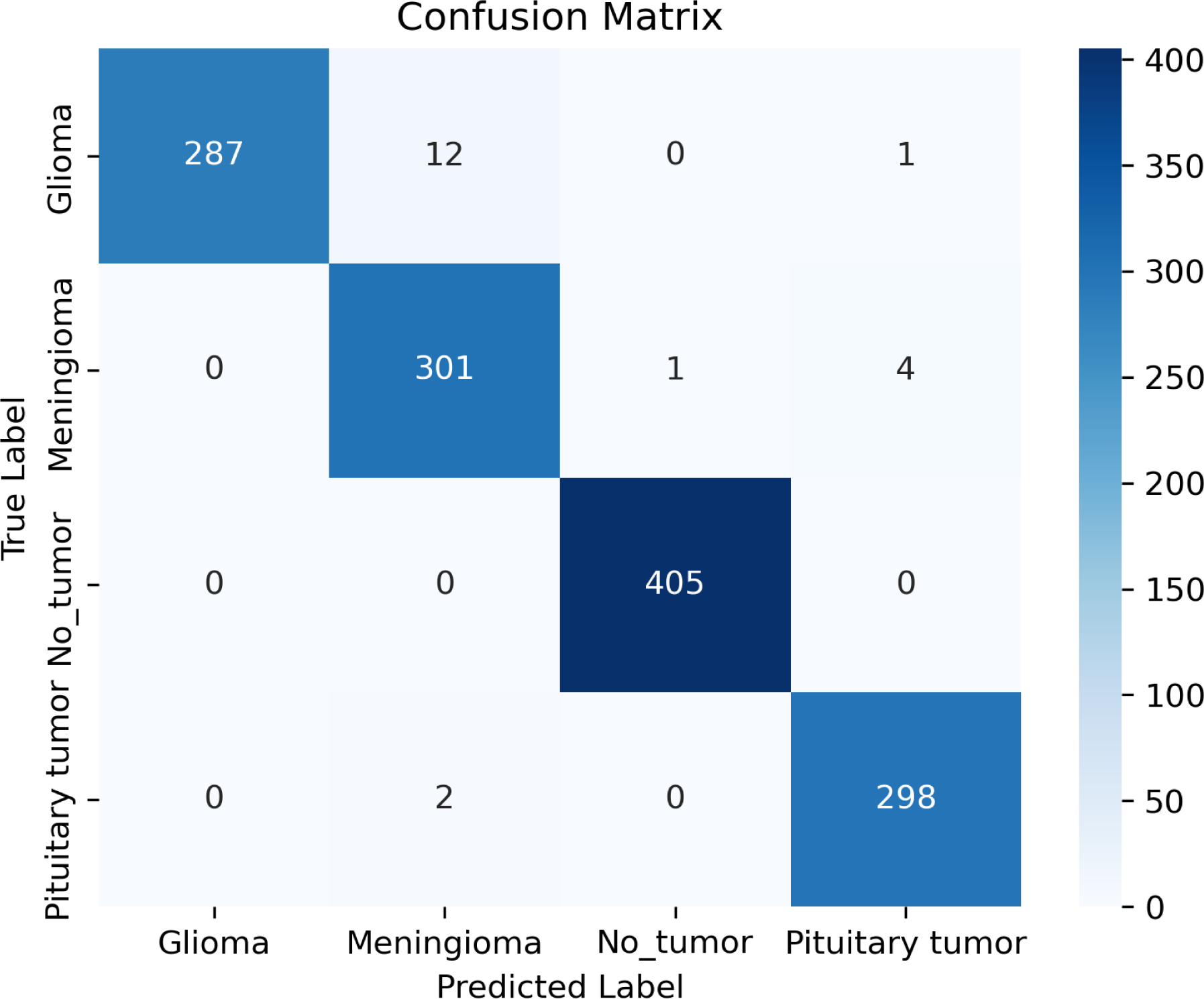
Confusion matrix

### Learning Rate Optimization

The learning rate schedule was strategically managed to optimize the training process (Figure 5). Initially set high for broader exploration, the learning rate was gradually decreased to refine the model’s convergence on optimal weights, facilitating robust learning without overfitting.

**Figure 5.**
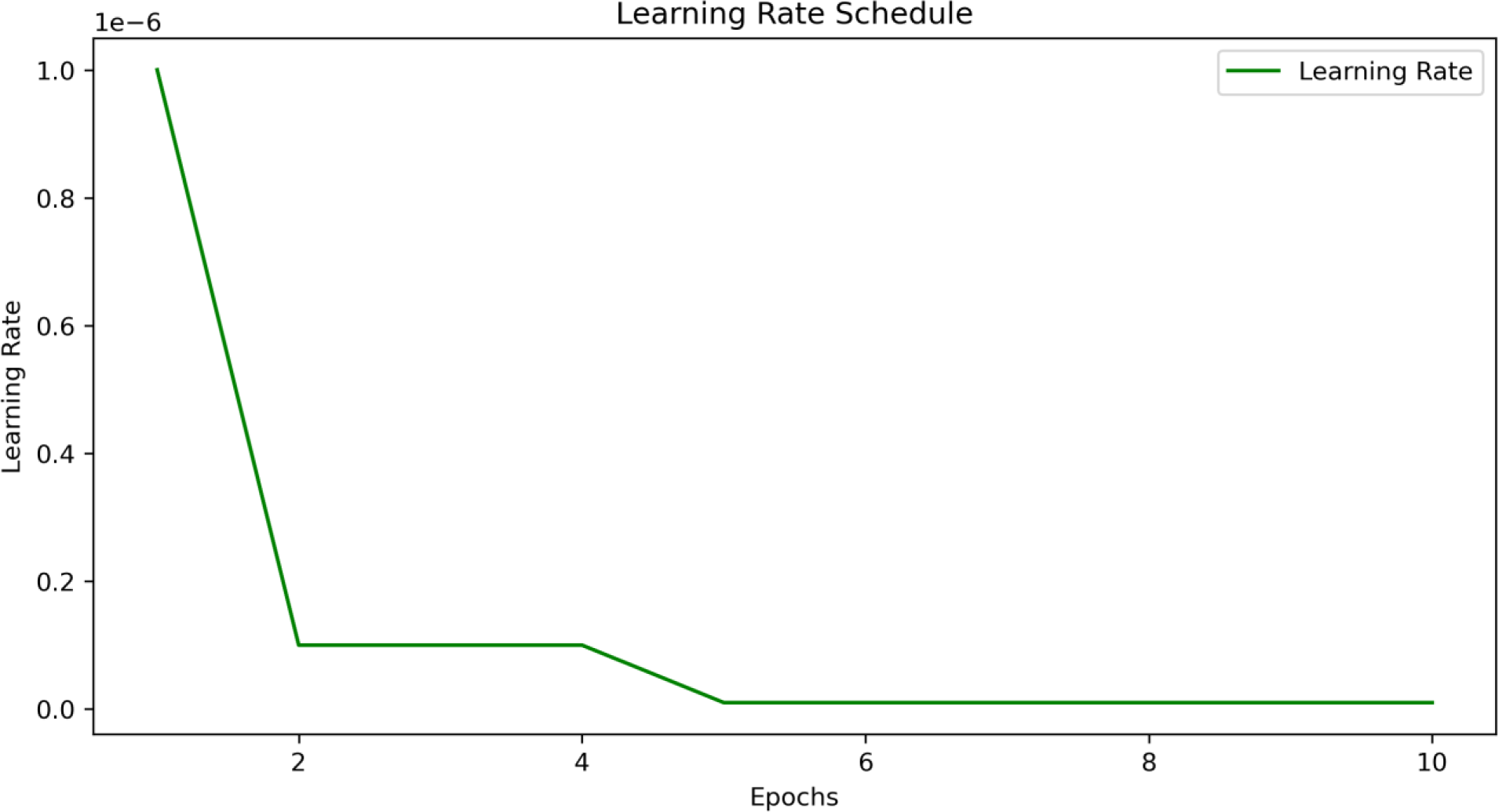
The learning rate schedule used during training. An optimized learning rate schedule was crucial in achieving efficient convergence and high model accuracy.

### Loss Metrics

Throughout the training epochs, the model exhibited a consistent decrease in training loss, while the validation loss remained low and stable, indicating a good model fit without overfitting (Figure 6). The convergence of training and validation loss suggests that the model was well-tuned to the complexity of the task, with loss metrics corroborating the high accuracy and AUC results.

**Figure 6.**
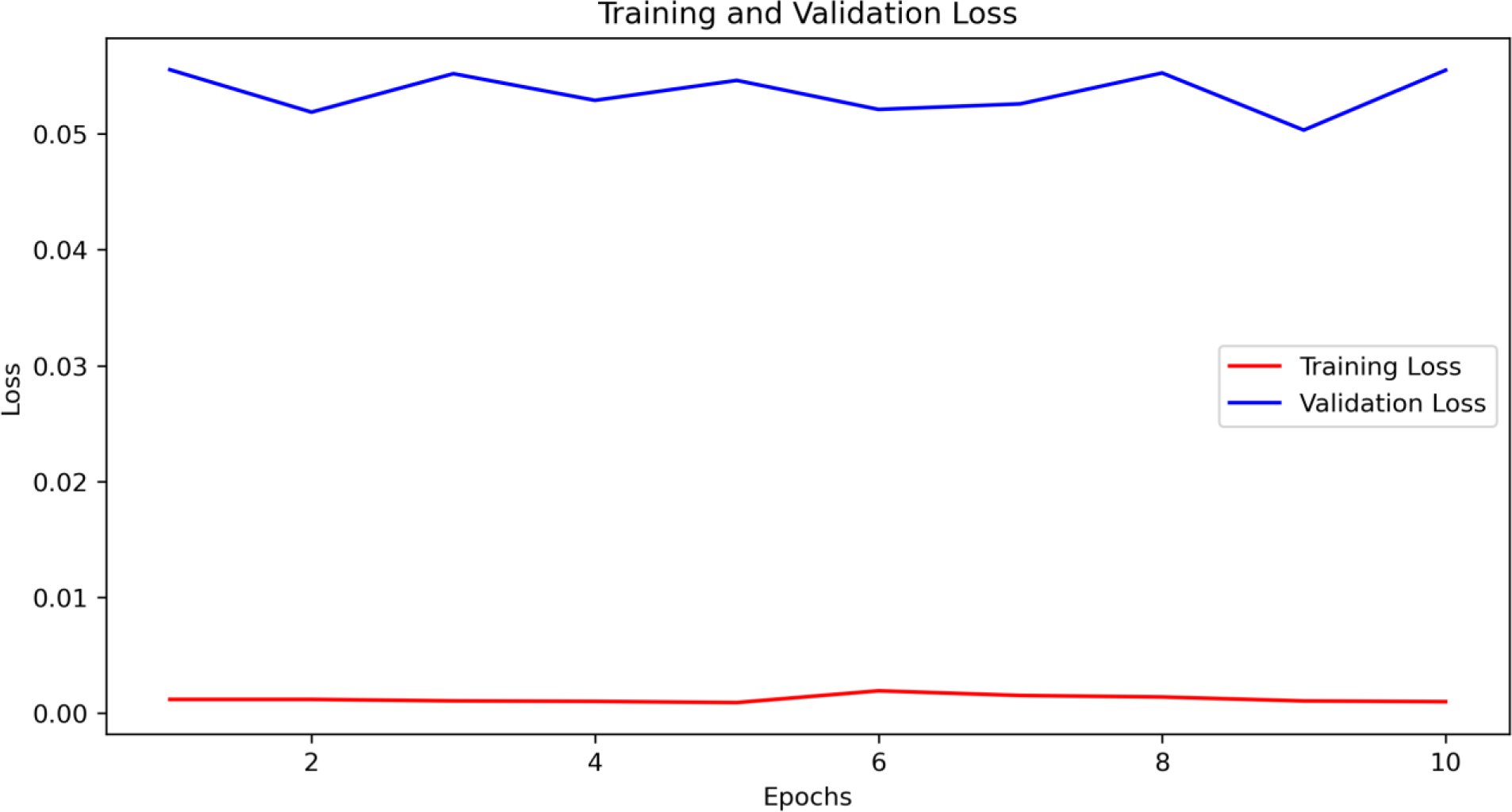
The training and validation loss curves over epochs. The decreasing loss values indicate effective model training and reduced overfitting, contributing to robust performance on unseen data.

### Discussion

In this study, we implemented a deep learning model for the detection of primary brain tumors using conventional MRI data. The model’s performance was assessed in practical scenarios, where it demonstrated robust capability to accurately predict brain tumor presence in unseen datasets. Impressively, it achieved a perfect AUC score of 1.0, reflecting its high sensitivity and precision in diagnosing both tumor and non-tumor MRI images. Recently, novel deep learning techniques have shown improved predictive performance in the medical domain, particularly in modeling complex and nonlinear effects compared to standard statistical prediction models. Hamd et al. constructed deep learning and machine learning models based on an artificial neural network (ANN), logistic regression, Gradient boosting, and random forest to predict glioma, permeative neuroectodermal, and pituitary tumors based on MRI images [8]. Hamd et al. concluded that ANN was superior to the rest models achieving an AUC of 0.97. In our study, we observed a higher AUC when using CNNs (AUC=1) compared to Hamd et al. traditional ANN (AUC=0.97) for the detection of glioma tumors. This improvement highlights the enhanced capability of CNNs in processing and analyzing complex medical images. This enhancement can be attributed to the inherent design of CNNs, which are optimized for image analysis by leveraging spatial hierarchies and local patterns within the images. Specifically, we employed the ResNet-18 model, a deep residual network architecture developed by He et al. [9], which was pre-trained on the ImageNet dataset comprising over 14 million images. The use of ResNet-18 allowed us to exploit its advanced feature extraction capabilities, resulting in more accurate and reliable detection of brain tumors compared to traditional ANN approaches. The higher AUC achieved by our CNN-based approach underscores the importance of using specialized deep-learning models like CNN with utilizing pre trained model such as ResNet-18 for medical image analysis. These models, pre-trained on extensive datasets, provide a robust framework for tackling complex medical imaging challenges, ultimately leading to improved diagnostic accuracy and better patient outcomes. “It’s important to note that while our study employed ResNet-18 for its efficiency, there are more complex models available, such as ResNet-34 and ResNet-50. Increasing the complexity of the model carries a risk of overfitting, particularly with datasets that already achieve high performance metrics like an AUC of 1. However, for datasets yielding lower performance, exploring these more advanced models could be beneficial to enhance diagnostic accuracy.

The reason we chose the CNN model for our model, was due to the CNN architecture is specifically designed to handle image data by leveraging layers that perform convolution operations to detect features such as edges, textures, and shapes within the images. These convolutional layers scan the MRI images with multiple filters to create feature maps that highlight important patterns. Pooling layers then reduce the dimensions of these feature maps, retaining critical information while minimizing computational load. Finally, fully connected layers combine these features to classify the images into their respective categories. This hierarchical feature extraction process allows CNNs to effectively capture the spatial and structural nuances of MRI scans, leading to accurate detection and differentiation of various brain tumor types. The ability of CNNs to automatically learn and extract relevant features from complex medical images makes them particularly suitable for medical image analysis, resulting in high diagnostic accuracy and reliability.

The application of AI in neuro-oncology has proven not only to enhance diagnostic accuracy but also to significantly improve efficiency. A notable competition involved a brain tumor diagnosis comparing the performances of human radiologists and an AI system developed by the Artificial Intelligence Research Centre for Neurological Disorders and Capital Medical University. In the competition the AI system, Biomind, achieved an 87% accuracy rate, diagnosing 195 out of 225 cases correctly within 15 minutes. In contrast, a team of 15 radiologists manually diagnosed 148 cases correctly, achieving a 66% accuracy rate over 30 minutes [10]. This demonstrates AI’s potential to revolutionize diagnostic processes in neuro-oncology by delivering faster and more accurate results.

Based on the confusion matrix, we observed that 12 MRI images of glioma were incorrectly predicted as meningioma. Patel et al. reported similar diagnostic challenges, describing two cases where MRI initially suggested meningiomas, but surgical outcomes confirmed glioblastomas [11]. They attributed these diagnostic errors to gliomas exhibiting MRI characteristics typically associated with meningiomas, such as the dural tail sign, CSF cleft sign, and broad dural contact. These imaging mimicry patterns could account for the misdiagnoses observed, suggesting that even with the application of AI, misdiagnosis is possible due to the inherent similarities between diseases. This highlights a limitation within AI applications in neuro-oncology that must be addressed in future studies. Additionally, it’s crucial to acknowledge that medical images often face issues like artifacts and resolution degradation, which can impact the accuracy of machine learning-based diagnostic methods.

The utilization of AI in neuro-oncology holds transformative potential by transitioning from exclusive reliance on radiologist expertise to standardized and efficient diagnostics accessible worldwide. This is facilitated by cloud-based platforms that provide virtual machines, making the primary requirement for utilizing the model just a basic computer. Wahl et al. highlight how expert systems like our deep learning model could enhance healthcare in resource-limited settings [12]. Such systems can assist physicians with diagnostics and treatment decisions, similar to practices in wealthier nations, and even substitute for human experts in areas where they are scarce [11]. This is particularly valuable in under-resourced communities. However, a significant challenge remains in these settings—the limited availability of MRI machines [13]. Addressing this issue requires a dual approach: integrating existing AI technologies to mitigate the absence of medical experts and increasing the accessibility of MRI technology in low-income countries.

Our study’s strengths include the use of a pre-trained ResNet-18 CNN, which provided high diagnostic accuracy and AUC in classifying glioma, meningioma, pituitary tumors, and non-tumor MRI images. Utilizing publicly available Kaggle datasets ensures replicability and transparency. However, limitations exist, such as the use of 2D MRI slices, and potentially missing critical volumetric data from 3D images. The Kaggle dataset’s lack of clinical diversity may affect generalizability. Furthermore, the lack of longitudinal data and the necessity for additional clinical validation on larger databases underscore areas requiring future research.

## Conclusions

Our study utilized a fine-tuned ResNet-18 model in PyTorch to significantly enhance the diagnostic accuracy of gliomas, meningiomas, and pituitary tumors. The model demonstrated high accuracy, indicating its potential to improve early diagnosis and patient outcomes. These findings suggest that integrating AI models like ResNet-18 into clinical workflows could lead to earlier detection and more precise prognostic evaluations, ultimately enhancing patient care and survival rates. Future research should focus on further validating these models with larger datasets and exploring their clinical applications.

## Data Availability

All data produced are available online at kaggle

https://doi.org/10.34740/KAGGLE/DSV/2645886

## Abbreviation

AI: Artificial Intelligence
AUC: Area Under the Curve
CNN: Convolutional Neural Network
CSF: Cerebrospinal Fluid
FLAIR: Fluid-Attenuated Inversion Recovery
IRB: Institutional Review Board
MRI: Magnetic Resonance Imaging
ROC: Receiver Operating Characteristic
ANN: Artificial Neural Network
ResNet-18: Residual Network 18 layer

## Author Contributions

Conceptualization, Alon G, Avner G; Methodology, Alon G, WS, TL,; Formal Analysis, Alon G, TL, WS; Writing – Original Draft Preparation, Alon G; Writing – Review & Editing, Alon G, Avner G; Supervision, Avner G

